# Occupation as a risk factor for progression of chronic kidney disease: retrospective cohort study

**DOI:** 10.1101/2021.03.28.21254487

**Authors:** Daisuke Takada, Susumu Kunisawa, Akira Kikuno, Tomoko Iritani, Yuichi Imanaka

## Abstract

**Background:** Several social determinants of health are associated with the progression of chronic kidney disease (CKD). We compared the risk of CKD progression among 18 occupational classifications using an annual health-checkup database.

**Methods:** We used the annual health checkup data and health insurance claims data of the Japan Health Insurance Association in Kyoto prefecture between April 2012 and March 2016. The primary outcome for survival analysis was defined as a more than 30% change in the estimated glomerular filtration rate (eGFR) from the first health checkup. We used the Cox proportional-hazards model for time-to-event analyses to estimate the hazard ratio, and 95% CIs for the primary outcome, adjusting for age, sex, eGFR, body mass index, blood pressure, blood sugar, dyslipidemia, uric acid, urinary protein, and existence of kidney diseases at first health checkup.

**Results:** We analyzed 239,506 employees, and 1,736 (0.7%) individuals whose eGFR had decreased by 30% or more; the mean follow-up period was 2.8 years (standard deviation: 1.2 years). When we compared the risk for “manufacturing,” five categories of industries (“information and communications”; “transport and postal services”; “accommodations, eating and drinking services”; “living-related and personal services and amusement service”; “medical, health care and welfare”) were associated with a decline in the risk of eGFR after adjusting for the confounding factors and/or mediators.

**Conclusions:** We provided evidence that the risk of CKD progression depends on occupation type. Further research is needed to confirm the mechanism and causal relationships involved.

## Introduction

Chronic kidney disease (CKD) is recognized as one of the most serious problems in public health. The global prevalence of CKD reached approximately 10% worldwide, affecting 500 million people. [1] Currently, the estimated glomerular filtration ratio (eGFR) is a good marker for kidney function, as a gradual reduction in eGFR leads to kidney failure in the general population. [2] Traditional lifestyle diseases such as diabetes and hypertension influence the development of CKD, increasing the risk of cardiovascular and cerebrovascular events, and some cancers. [1, 3]

Recently, the etiology of non-traditional disorders has gained attention, including novel risk factors for CKD. Systematic reviews have shown that occupational status, including agricultural and livestock-related work, is associated with an increased risk of CKD progression. [4-8] The causal role of occupation in renal dysfunction might be accounted for by several mechanisms, including a greater prevalence of hyperphosphatemia [9], work-related heat stress [10], and lower health-related quality of life both mentally and physically. [11, 12] However, this field of study is only just emerging, and more studies are needed to clarify such associations. Here, we report the association between occupation and the progression of CKD through an extensive health-checkup database in Japan.

## Methods

### Database, medical records and target populations

We conducted a retrospective database analysis using annual health checkup data between April 2012 and March 2016 and health insurance claims data, together with information on the subject’s employer, from a database of the Japan Health Insurance Association in Kyoto prefecture, Japan[13]. Annual specific medical checkups for employees aged ≥ 40 years were established in the following law: “Act on Assurance of Medical Care for Elderly People”. Accordingly, the Japan Health Insurance Association get employees aged ≥ 35 years to have annual health checkups until they lose their eligibility, such as their changing jobs, being fired, or passed away. We enrolled employees who were aged more than 35 years of age and had undergone two or more health checkups during the observation period. In our analysis, we excluded participants: 1) Whose eGFR was less than 30 mL/min/1.73m^2^; 2) who had several kidney diseases; and 3) those who were missing data from their first health checkup. Kidney diseases were defined using ICD-10 codes recorded in the claims data, such as glomerular disease, renal artery stenosis, and polycystic kidney disease (N00-08, I70, and Q61).

### Covariates

Covariateds include patient sex, age, body mass index (BMI), eGFR, urinary protein, abdominal circumference, blood pressure (BP), dyslipidemia, diabetes, uric acid and kidney disease. All the covariates were collected at the first health checkup, and eGFR was calculated by the formula for Japanese patients devised by Matsuo et al. [14]

The patients were classified as follows: four groups based on age (35–44, 45–54, 55–64, and ≥65 years); four groups based on BMI (<18.5 kg/m^2^, thin; ≥18.5 and <25 kg/m^2^, normal range; ≥25 and <30 kg/m^2^, pre-obesity; and ≥30 kg/m^2^, obesity) according to the World Health Organization [15]; five groups based on eGFR (>60, 60–45, 45–30 mL/min/1.73m^2^); three groups based on urinary protein (using dipsticks ≥ 1+ (positive), ±(trace), and negative), abdominal circumference (if male ≥ 85 cm, female ≥ 90 cm); five groups based on BP (systolic blood pressure (sbp) ≥180 mmHg or diastolic blood pressure (dbp) ≥110 mmHg without antihypertensives, sbp ≥160 mmHg or dbp ≥100 mmHg without drugs, sbp ≥140 mmHg or dbp ≥90 mmHg without drugs, normal without drugs, and with drugs); and four groups based on dyslipidemia (triglyceride ≥150 mg/dL or high-density lipoprotein cholesterol <40 mg/dL defined as abnormality with hypolipidemic drugs, abnormality without hypolipidemic drugs, normal with hypolipidemic drugs, normal without hypolipidemic drugs), diabetes (fasting blood sugar ≥110 mg/dL or hemoglobin A1c ≥5.6% defined as abnormality with antidiabetic drugs, abnormality without antidiabetic drugs, normal with antidiabetic drugs, normal without antidiabetic drugs), and hyperuricemia (defined uric acid ≥8 mg/dl without anti-hyperuricemias or using anti-hyperuricemias).

Occupational information was acquired from notifications when companies were established and submitted information to the Japan Pension Service, so that their employees were covered by health insurance. The occupational information was classified into 18 categories according to the Japan Standard Industrial Classification, as modified from the International Standard Industrial Classifications of All Economic Activities. The classifications are as follows: 1) agriculture and forestry and fishers, 2) mining and quarrying of stone, 3) construction, 4) manufacturing, 5) electricity, gas, heat supply and water production, transmission and distribution of electricity, 6) information and communications, 7) transport and postal services, 8) wholesale and retail trace, 9) finance and insurance, 10) real estate and goods rental and leasing, 11) scientific research, professional and technical service, 12) accommodations, eating and drinking services, 13) living-related and personal services and amusement service, 14) education, learning support school education, 15) medical, health care and welfare, 16) compound services, such as establishments engaged in administrative or ancillary economic activities, 17) other services, such as waste disposal business, 18) government, except where elsewhere classified as national government service. The primary industry is classified as category “1”, the secondary industry is classified as category “2” to “4”, and the tertiary industry is classified as category “5” to “18”.

### Statistical analysis

The primary outcome for the survival analysis per person-year was defined as a decrease of 30% or more in eGFR from the baseline eGFR at the time of their first medical check-up. [2] The eGFR was calculated by the equation used by the Japanese Society of Nephrology. [14] Patients were followed until there was a decline in their eGFR of at least 30% (event), or censored due to death, a change in their insurance contract, or the date of their last health check-up, whichever came first.

The Cox proportional-hazards model was used for time-to-event analyses to estimate the hazard ratio; 95% CIs were calculated for the primary outcome. Follow-up period data for patients were censored on the date of the last health checkup. The analysis used two types of models: Model 1, adjusted for age, sex, and eGFR at baseline (without medication factor); and Model 2, adjusted for age, sex, eGFR, body mass index, blood pressure, blood sugar, dyslipidemia, uric acid, and urinary protein at baseline (with medication factor).

Subgroup analysis was performed in Model 2, where the analysis population was limited according to eGFR from G1 to G2 (eGFR >60 mL/min/1.73m^2^).

### Ethical considerations

The research protocol was approved by the ethics committee of the Kyoto University Graduate School and Faculty of Medicine (approval number: R1631) and by the ethics committee of the Japan Health Insurance Association. The participants were informed to use their data in epidemiological studies that dealt with personal information in a manner that was not identifiable, for example, we cannot show less than 10 people in the tables. Due to the law: “Act on the Protection of Personal Information”, the data cannot be released to the public. The data will be shared on reasonable request to the corresponding author.

## Results

A total of 253,673 employees were enrolled and fulfilled the inclusion criteria; 14,218 (5.6%) were excluded due to missing data. We analyzed the remaining 237,987 employees (Fig 1). By the end of follow-up period, there were 1,736 (0.7%) individuals whose eGFR had decreased by 30% or more; the mean follow-up period was 2.8 years (standard deviation: 1.2 years).

**Figure 1:**
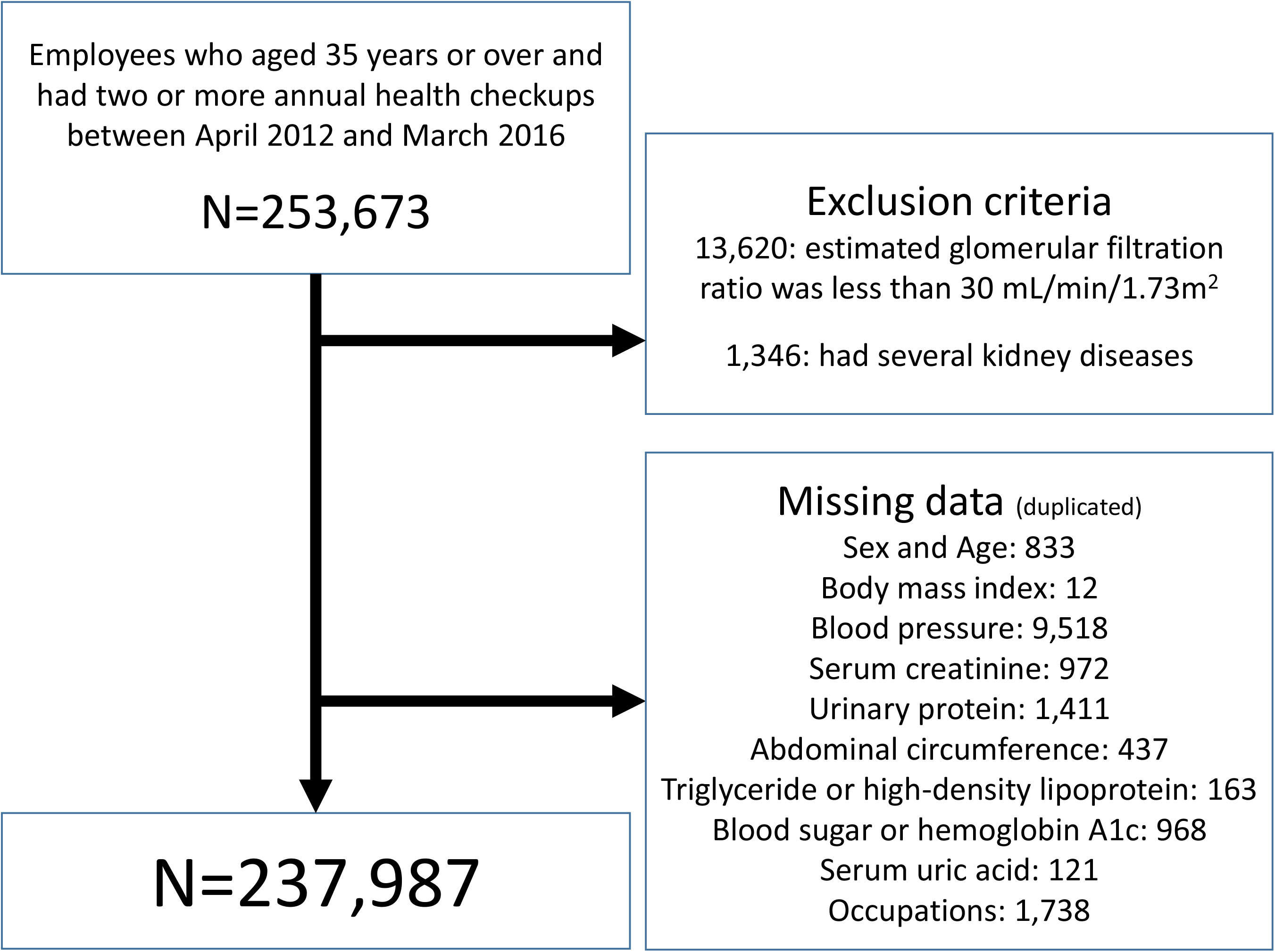
Flow chart of patient selection N: number of patients

The baseline characteristics of the patients are shown in Table 1. As manufacturing was the most popular category, we used manufacturing as the reference category.

**Table 1.**
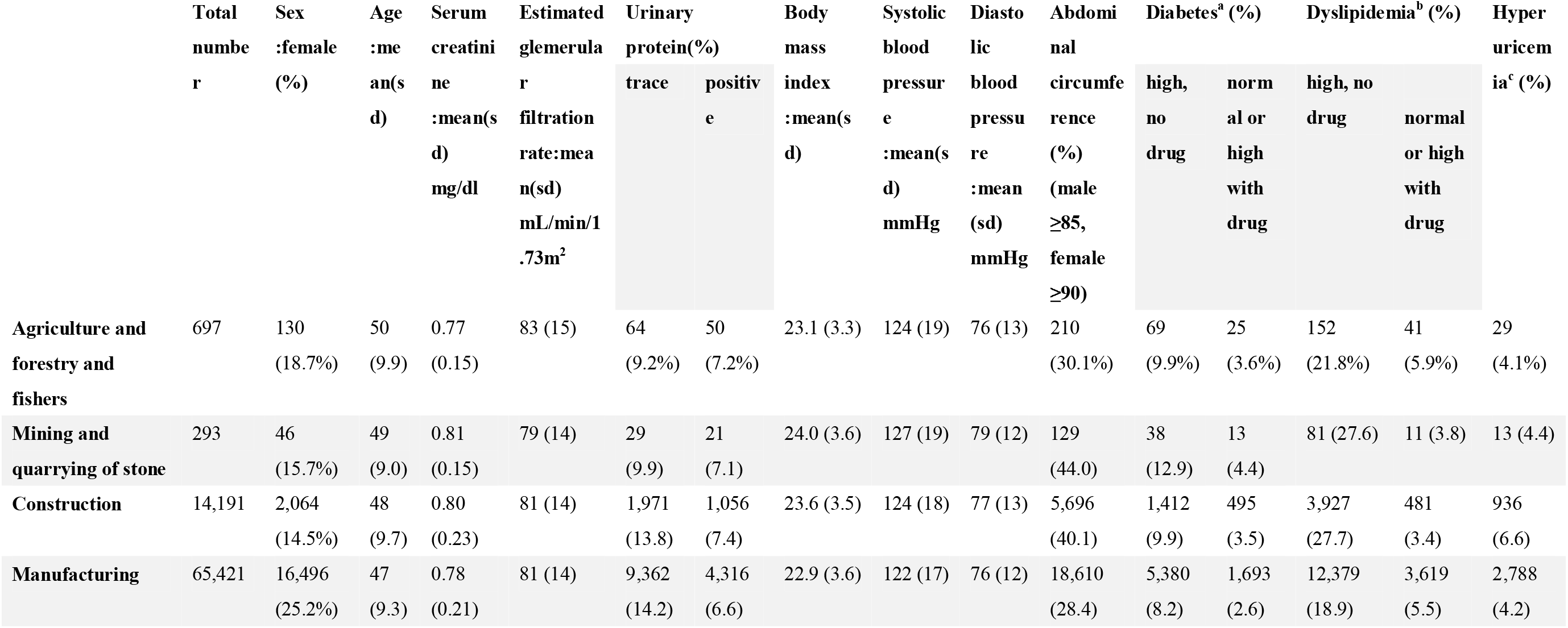
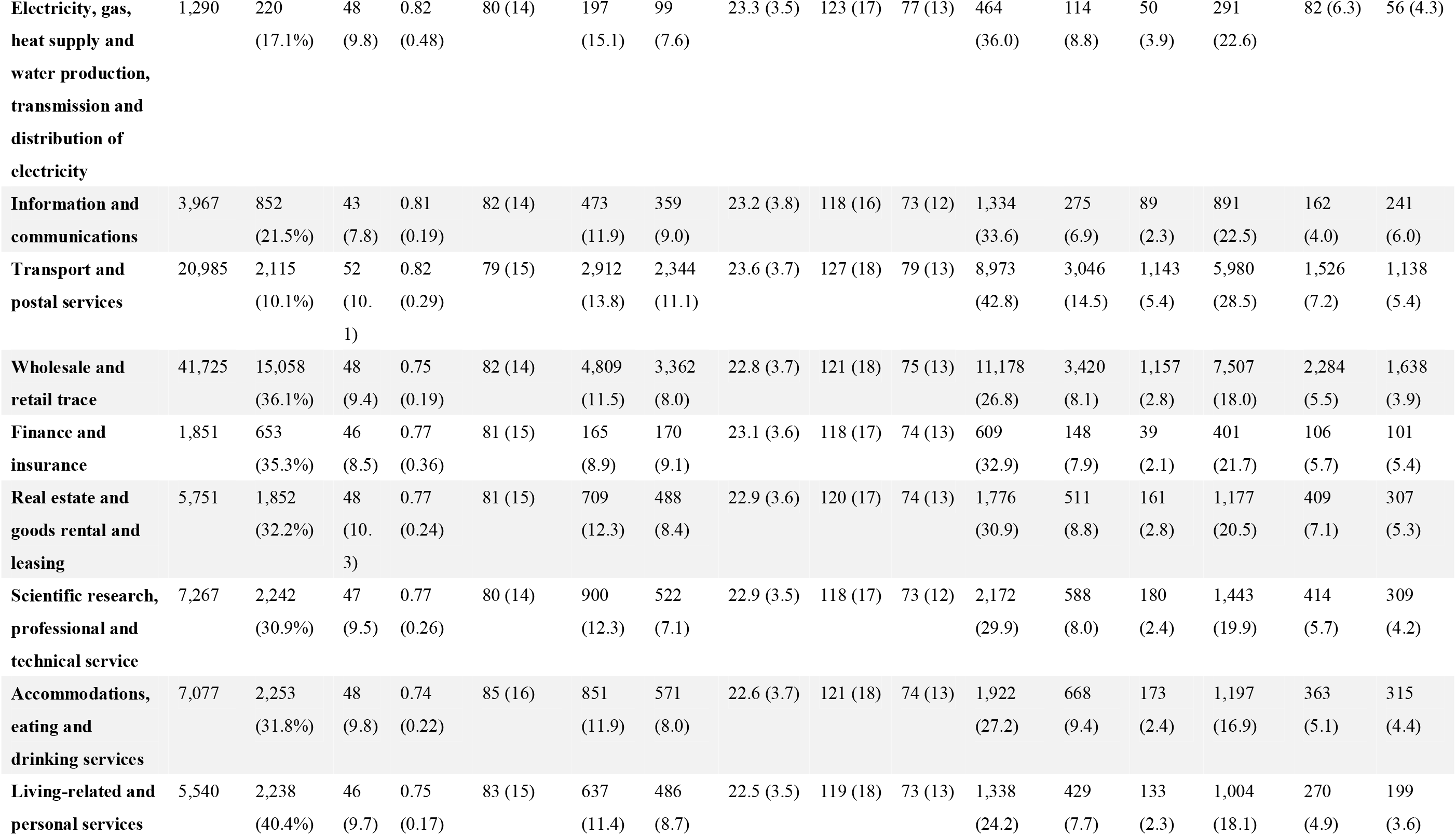
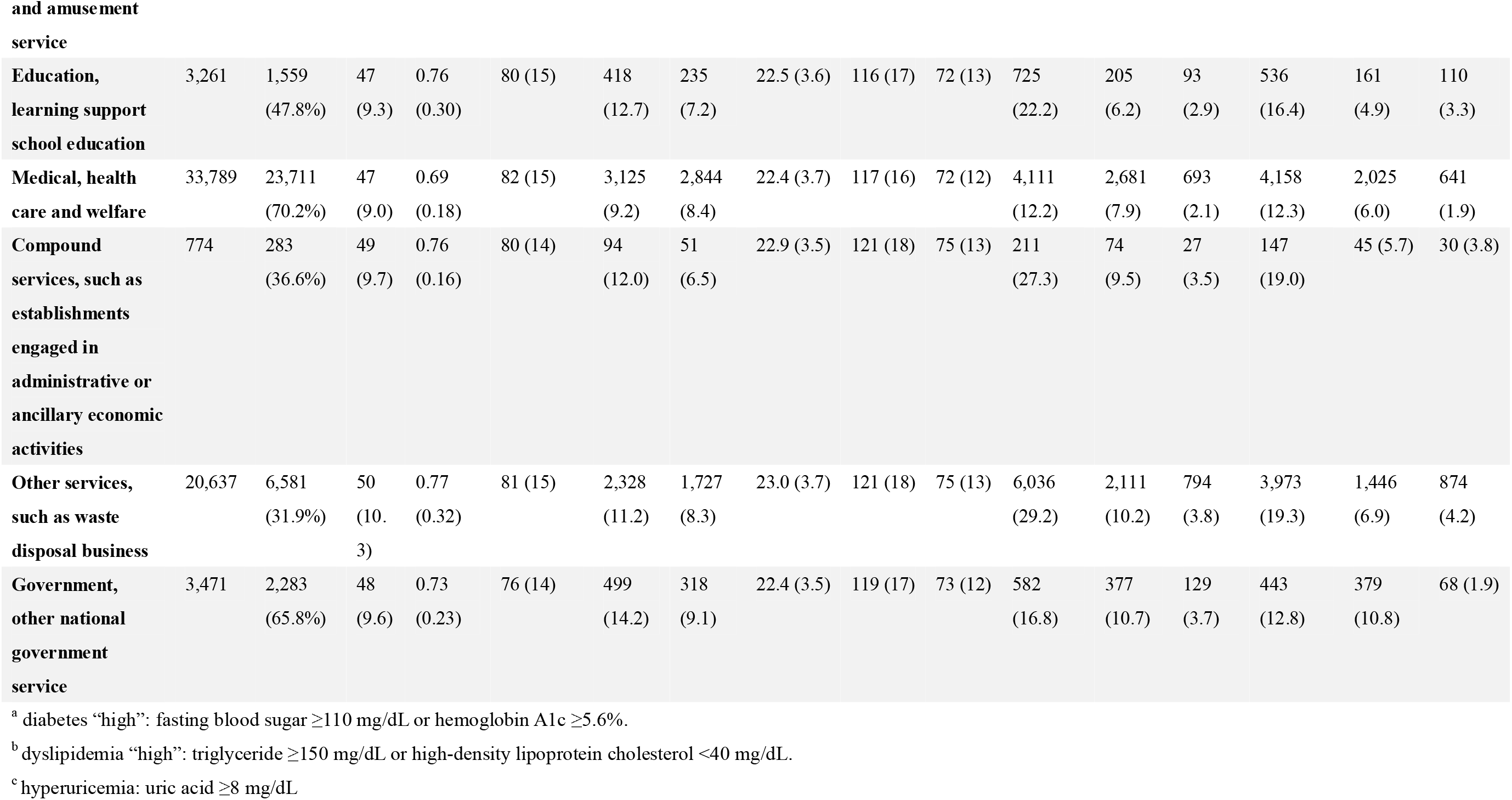
selected baseline characteristics according to the occupational information categories

|  | Total<br>number | Sex<br>:female<br>(%) | Age<br>:mean(s<br>d) | Serum<br>creatinine<br>:mean(s<br>d)<br>mg/dl | Estimated<br>glomerular<br>filtration<br>rate:mean<br>(sd)<br>mL/min/1<br>.73m<sup>2</sup> | Urinary<br>protein(%)<br>trace positive |  | Body<br>mass<br>index<br>:mean(s<br>d) | Systolic<br>blood<br>pressure<br>:mean(s<br>d)<br>mmHg | Diastolic<br>blood<br>pressure<br>:mean<br>(sd)<br>mmHg | Abdominal<br>circumference<br>(%)(male<br>≥85,<br>female<br>≥90) | Diabetes<sup>a</sup> (%)<br>high, no drug<br>normal or high<br>with drug |  | Dyslipidemia<sup>b</sup> (%)<br>high, no drug<br>normal or high<br>with drug |  | Hyperuricemia<sup>c</sup> (%) |
| --- | --- | --- | --- | --- | --- | --- | --- | --- | --- | --- | --- | --- | --- | --- | --- | --- |
| **Agriculture and forestry and fishers** | 697 | 130<br>(18.7%) | 50<br>(9.9) | 0.77<br>(0.15) | 83 (15) | 64<br>(9.2%) | 50<br>(7.2%) | 23.1 (3.3) | 124 (19) | 76 (13) | 210<br>(30.1%) | 69<br>(9.9%) | 25<br>(3.6%) | 152<br>(21.8%) | 41<br>(5.9%) | 29<br>(4.1%) |
| **Mining and quarrying of stone** | 293 | 46<br>(15.7%) | 49<br>(9.0) | 0.81<br>(0.15) | 79 (14) | 29<br>(9.9) | 21<br>(7.1) | 24.0 (3.6) | 127 (19) | 79 (12) | 129<br>(44.0) | 38<br>(12.9) | 13<br>(4.4) | 81 (27.6) | 11 (3.8) | 13 (4.4) |
| **Construction** | 14,191 | 2,064<br>(14.5%) | 48<br>(9.7) | 0.80<br>(0.23) | 81 (14) | 1,971<br>(13.8) | 1,056<br>(7.4) | 23.6 (3.5) | 124 (18) | 77 (13) | 5,696<br>(40.1) | 1,412<br>(9.9) | 495<br>(3.5) | 3,927<br>(27.7) | 481<br>(3.4) | 936<br>(6.6) |
| **Manufacturing** | 65,421 | 16,496<br>(25.2%) | 47<br>(9.3) | 0.78<br>(0.21) | 81 (14) | 9,362<br>(14.2) | 4,316<br>(6.6) | 22.9 (3.6) | 122 (17) | 76 (12) | 18,610<br>(28.4) | 5,380<br>(8.2) | 1,693<br>(2.6) | 12,379<br>(18.9) | 3,619<br>(5.5) | 2,788<br>(4.2) |
| **Electricity, gas, heat supply and water production, transmission and distribution of electricity** | 1,290 | 220<br>(17.1%) | 48<br>(9.8) | 0.82<br>(0.48) | 80 (14) | 197<br>(15.1) | 99<br>(7.6) | 23.3 (3.5) | 123 (17) | 77 (13) | 464<br>(36.0) | 114<br>(8.8) | 50<br>(3.9) | 291<br>(22.6) | 82 (6.3) | 56 (4.3) |
| **Information and communications** | 3,967 | 852<br>(21.5%) | 43<br>(7.8) | 0.81<br>(0.19) | 82 (14) | 473<br>(11.9) | 359<br>(9.0) | 23.2 (3.8) | 118 (16) | 73 (12) | 1,334<br>(33.6) | 275<br>(6.9) | 89<br>(2.3) | 891<br>(22.5) | 162<br>(4.0) | 241<br>(6.0) |
| **Transport and postal services** | 20,985 | 2,115<br>(10.1%) | 52<br>(10.1) | 0.82<br>(0.29) | 79 (15) | 2,912<br>(13.8) | 2,344<br>(11.1) | 23.6 (3.7) | 127 (18) | 79 (13) | 8,973<br>(42.8) | 3,046<br>(14.5) | 1,143<br>(5.4) | 5,980<br>(28.5) | 1,526<br>(7.2) | 1,138<br>(5.4) |
| **Wholesale and retail trade** | 41,725 | 15,058<br>(36.1%) | 48<br>(9.4) | 0.75<br>(0.19) | 82 (14) | 4,809<br>(11.5) | 3,362<br>(8.0) | 22.8 (3.7) | 121 (18) | 75 (13) | 11,178<br>(26.8) | 3,420<br>(8.1) | 1,157<br>(2.8) | 7,507<br>(18.0) | 2,284<br>(5.5) | 1,638<br>(3.9) |
| **Finance and insurance** | 1,851 | 653<br>(35.3%) | 46<br>(8.5) | 0.77<br>(0.36) | 81 (15) | 165<br>(8.9) | 170<br>(9.1) | 23.1 (3.6) | 118 (17) | 74 (13) | 609<br>(32.9) | 148<br>(7.9) | 39<br>(2.1) | 401<br>(21.7) | 106<br>(5.7) | 101<br>(5.4) |
| **Real estate and goods rental and leasing** | 5,751 | 1,852<br>(32.2%) | 48<br>(10.3) | 0.77<br>(0.24) | 81 (15) | 709<br>(12.3) | 488<br>(8.4) | 22.9 (3.6) | 120 (17) | 74 (13) | 1,776<br>(30.9) | 511<br>(8.8) | 161<br>(2.8) | 1,177<br>(20.5) | 409<br>(7.1) | 307<br>(5.3) |
| **Scientific research, professional and technical service** | 7,267 | 2,242<br>(30.9%) | 47<br>(9.5) | 0.77<br>(0.26) | 80 (14) | 900<br>(12.3) | 522<br>(7.1) | 22.9 (3.5) | 118 (17) | 73 (12) | 2,172<br>(29.9) | 588<br>(8.0) | 180<br>(2.4) | 1,443<br>(19.9) | 414<br>(5.7) | 309<br>(4.2) |
| **Accommodations, eating and drinking services** | 7,077 | 2,253<br>(31.8%) | 48<br>(9.8) | 0.74<br>(0.22) | 85 (16) | 851<br>(11.9) | 571<br>(8.0) | 22.6 (3.7) | 121 (18) | 74 (13) | 1,922<br>(27.2) | 668<br>(9.4) | 173<br>(2.4) | 1,197<br>(16.9) | 363<br>(5.1) | 315<br>(4.4) |
| **Living-related and personal services** | 5,540 | 2,238<br>(40.4%) | 46<br>(9.7) | 0.75<br>(0.17) | 83 (15) | 637<br>(11.4) | 486<br>(8.7) | 22.5 (3.5) | 119 (18) | 73 (13) | 1,338<br>(24.2) | 429<br>(7.7) | 133<br>(2.3) | 1,004<br>(18.1) | 270<br>(4.9) | 199<br>(3.6) |
| **Education,**<br>**learning support**<br>**school education** | 3,261 | 1,559<br>(47.8%) | 47<br>(9.3) | 0.76<br>(0.30) | 80 (15) | 418<br>(12.7) | 235<br>(7.2) | 22.5 (3.6) | 116 (17) | 72 (13) | 725<br>(22.2) | 205<br>(6.2) | 93<br>(2.9) | 536<br>(16.4) | 161<br>(4.9) | 110<br>(3.3) |
| **Medical, health**<br>**care and welfare** | 33,789 | 23,711<br>(70.2%) | 47<br>(9.0) | 0.69<br>(0.18) | 82 (15) | 3,125<br>(9.2) | 2,844<br>(8.4) | 22.4 (3.7) | 117 (16) | 72 (12) | 4,111<br>(12.2) | 2,681<br>(7.9) | 693<br>(2.1) | 4,158<br>(12.3) | 2,025<br>(6.0) | 641<br>(1.9) |
| **Compound**<br>**services, such as**<br>**establishments**<br>**engaged in**<br>**administrative or**<br>**ancillary economic**<br>**activities** | 774 | 283<br>(36.6%) | 49<br>(9.7) | 0.76<br>(0.16) | 80 (14) | 94<br>(12.0) | 51<br>(6.5) | 22.9 (3.5) | 121 (18) | 75 (13) | 211<br>(27.3) | 74<br>(9.5) | 27<br>(3.5) | 147<br>(19.0) | 45 (5.7) | 30 (3.8) |
| **Other services,**<br>**such as waste**<br>**disposal business** | 20,637 | 6,581<br>(31.9%) | 50<br>(10.3) | 0.77<br>(0.32) | 81 (15) | 2,328<br>(11.2) | 1,727<br>(8.3) | 23.0 (3.7) | 121 (18) | 75 (13) | 6,036<br>(29.2) | 2,111<br>(10.2) | 794<br>(3.8) | 3,973<br>(19.3) | 1,446<br>(6.9) | 874<br>(4.2) |
| **Government,**<br>**other national**<br>**government**<br>**service** | 3,471 | 2,283<br>(65.8%) | 48<br>(9.6) | 0.73<br>(0.23) | 76 (14) | 499<br>(14.2) | 318<br>(9.1) | 22.4 (3.5) | 119 (17) | 73 (12) | 582<br>(16.8) | 377<br>(10.7) | 129<br>(3.7) | 443<br>(12.8) | 379<br>(10.8) | 68 (1.9) |

<sup>a</sup> diabetes “high”: fasting blood sugar ≥110 mg/dL or hemoglobin A1c ≥5.6%. <sup>b</sup> dyslipidemia “high”: triglyceride ≥150 mg/dL or high-density lipoprotein cholesterol <40 mg/dL. <sup>c</sup> hyperuricemia: uric acid ≥8 mg/dL

Cox regression analysis was performed to identify the predictors of renal survival in the two models (Table 2). After adjusting for all the confounding factors, five categories of industries were associated with the risk of eGFR decrease. The hazard ratios and 95% confidence estimated in Model 2 were 1.69 (1.18-2.42) for “information and communications”; 1.37 (1.15-1.64) for “transport and postal services”; 1.55 (1.18-2.04) for “accommodations, eating and drinking services”; 1.44 (1.06-1.97) for “Living-related and personal services and amusement service”; 1.56 (1.34-1.82) for “medical, health care and welfare”.

**Table 2.** The incident rate per 1000 person-years and hazard ratios for a decrease of 30% or more in estimated glomerular filtration ratio in crude and adjusted models

| Sector of industry | Japan Standard Industrial Classification | Number | Incident rate per 1000 person-years (95% CI) | Model 1<sup>a</sup> | Model 2<sup>b</sup> |
| --- | --- | --- | --- | --- | --- |
| Primary industry | 1) Agriculture and forestry and fishers | 697 | 4.1 (1.9–7.5) | 1.93 (0.96–3.88) | 1.90 (0.94–3.83) |
| Secondary industry | 2) Mining and quarrying of stone | 293 | 2.1 (0.4–6.6) | 1.03 (0.26–4.14) | 0.88 (0.22–3.53) |
|  | 3) Construction | 14,191 | 2.5 (2.1–3.1) | **1.29\* (1.04–1.60)** | 1.23 (0.99–1.52) |
|  | 4) Manufacturing | 65,421 | 2.0 (1.8–2.2) | 1.0 (ref) | 1.0 (ref) |
| Tertiary industry | 5) Electricity, gas, heat supply and water production, transmission and distribution of electricity | 1,290 | 2.1 (1.5–5.0) | 1.51 (0.83–2.75) | 1.51 (0.83–2.75) |
|  | 6) Information and communications | 3,967 | 3.0 (2.0–4.1) | **1.74\* (1.21–2.50)** | **1.69\* (1.18–2.42)** |
|  | 7) Transport and postal services | 20,985 | 3.4 (3.0–3.9) | **1.60\* (1.34–1.90)** | **1.37\* (1.15–1.64)** |
|  | 8) Wholesale and retail trade | 41,725 | 2.3 (2.0–2.6) | 1.12 (0.96–1.31) | 1.10 (0.94–1.28) |
|  | 9) Finance and insurance | 1,851 | 2.0 (0.9–3.8) | 1.09 (0.54–2.19) | 1.06 (0.53–2.14) |
|  | 10) Real estate and goods rental and leasing | 5,751 | 2.6 (1.9–3.2) | 1.25 (0.91–1.73) | 1.25 (0.91–1.73) |
|  | 11) Scientific research, professional and technical service | 7,267 | 2.5 (1.9–3.2) | 1.24 (0.92–1.66) | 1.25 (0.93–1.67) |
|  | 12) Accommodations, eating and drinking services | 7,077 | 3.3 (2.5–4.2) | **1.59\* (1.21–2.09)** | **1.55\* (1.18–2.04)** |
|  | 13) Living-related and personal services and amusement service | 5,540 | 3.0 (2.2–4.0) | **1.48\* (1.08–2.01)** | **1.44\* (1.06–1.97)** |
|  | 14) Education, learning support school education | 3,261 | 1.6 (0.7–2.6) | 0.78 (0.47–1.31) | 0.77 (0.46–1.29) |
|  | 15) Medical, health care and welfare | 33,789 | 3.6 (3.2–4.0) | **1.59\* (1.37–1.86)** | **1.56\* (1.34–1.82)** |
|  | 16) Compound services, such as establishments engaged in administrative or ancillary economic activities | 774 | 2.7 (1.1–5.5) | 1.28 (0.57–2.86) | 1.25 (0.56–2.81) |
|  | 17) Other services, such as waste disposal business | 20,637 | 2.7 (2.3–3.1) | **1.21\* (1.0–1.47)** | 1.18 (0.98–1.43) |
|  | 18) Government, other national government service | 3,471 | 2.5 (1.6–3.7) | 0.95 (0.63–1.53) | 0.97 (0.62–1.50) |

Hazard ratio (95% confidence interval)\* in bold showed p-value <0.05 Abbreviation eGFR, estimated glomerular filtration rate; CI, confidence interval <sup>a</sup> Adjusted for sex, age and estimated glomerular filtration ratio at first health checkup <sup>b</sup> Adjusted for sex, age, body mass index, estimated glomerular filtration ratio, urinary protein, abdominal circumference, blood pressure, dyslipidemia, diabetes, uric acid and kidney disease at first health checkup

The other hazard ratios for a decrease of 30% or more in eGFR in Model 2 were shown in Figure 2.

**Figure 2:**
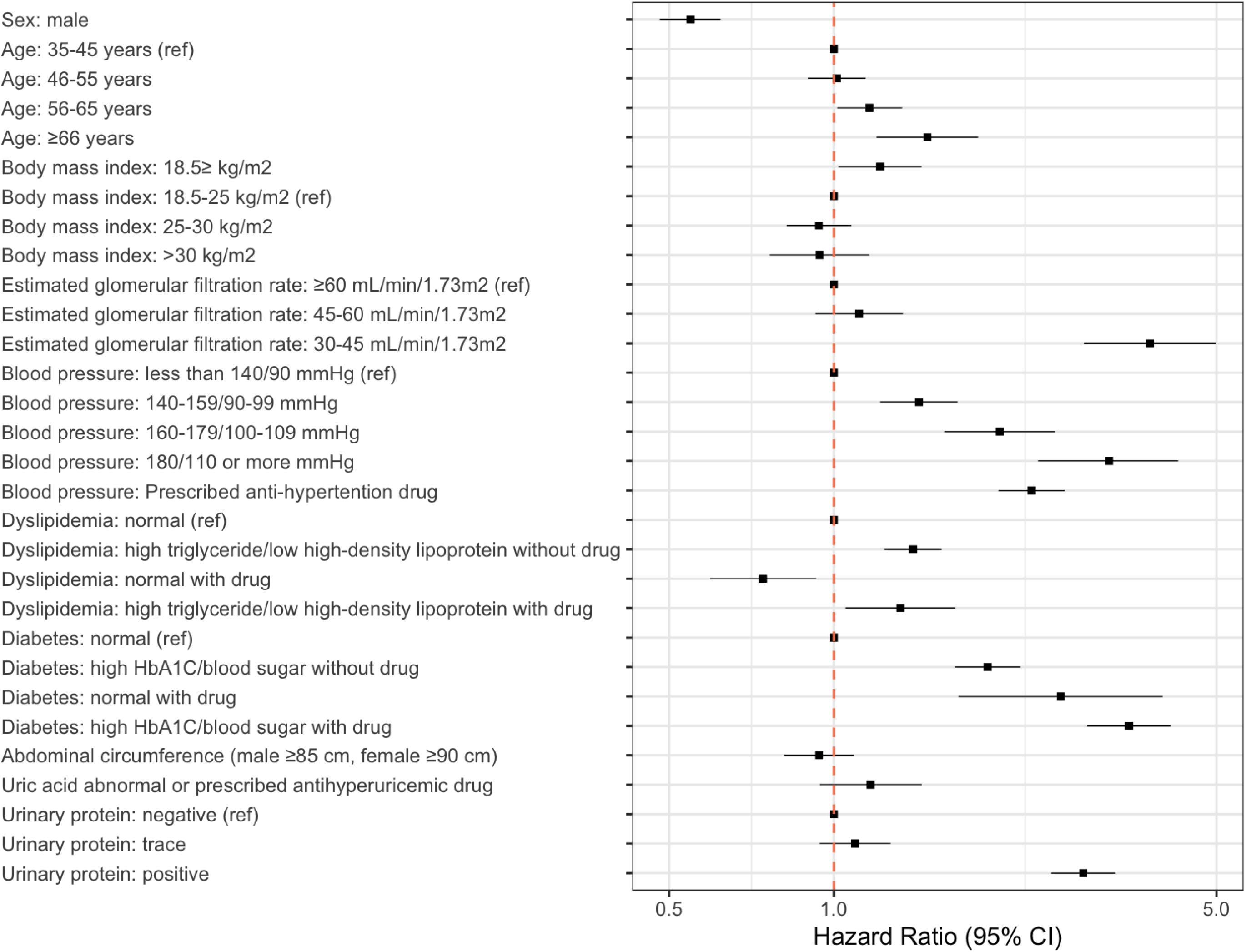
The other hazard ratios for a decrease of 30% or more in estimated glomerular filtration ratio, adjusted for sex, age, body mass index, estimated glomerular filtration ratio, urinary protein, abdominal circumference, blood pressure, dyslipidemia, diabetes, uric acid and kidney disease at first health checkup

The results for the subgroup analysis were similar to those for the main analysis.

## Discussion

We showed that type of occupation substantially influences CKD progression. In particular, when we compared risks to the risk of “manufacturing,” we found that five categories of industries (“Information and communications”; “Transport and postal services”; “Accommodations, eating and drinking services”; “Living-related and personal services and amusement service”; “Medical, health care and welfare”) were significantly associated with the risk of eGFR decline after adjusting for confounding factors and/or other mediators such as medication factors. Then we can interpret that these five categories include other risk factors than such medication factors.

Interestingly, we found that these categories share two working conditions: nonstandard work schedule and sedentary time. In the case of nonstandard work schedule, night shift work and rotating shift work may influence renal function by disrupting the endogenous circadian system with several hormonal and metabolic disturbances and negatively impacting physical and mental health. [16, 17] In fact, nonstandard work schedule is likely a risk factor for several vascular diseases [18, 19], and might result in hospitalization for cardiovascular disease, metabolic syndrome or various kinds of cancer. [16] Similarly, shift works has been shown to increase the risk of CKD onset in Japanese workers. [20] Thus, the four identified types of industry may decrease an employee’s renal function based on the employee’s nonstandard work schedule.

The other condition, sedentary time, is a feature of occupation categories such as “transport and postal services” and “information and communications,” both of which require people to sit long hours each day. Recently, several researchers have reported that prolonged sedentary time is associated with diseases such as diabetes and cardiovascular events and leads to an increase in total mortality [21-24], although the mechanism remains unknown. Likewise, prolonged sedentary time has also been associated with higher mortality due to kidney diseases. [25] Remarkably, we could change these conditions by changing the way to work, future researches need to elucidate the causal relationships.

According to a previous meta-analysis, agriculture is also related to kidney damage. [4] The authors of the study raised the possibility that the cause could be agrochemicals or sugar cane. The present study showed that agriculture, including forestry and fishers, significantly increased the risk of CKD progression in Model 1 but not in Model 2, suggesting that lifestyle disease, rather than occupation, could have an impact on the renal function in Japanese agricultural workers.

There were some limitations to our study. First, competing events, including office relocation and employees’ events, were regarded as censored in our analysis, and some of these might result in a healthy-worker bias. For example, the decline in an employee’s eGFR might cause other complications that result in them result in their quitting jobs, changing their jobs, or dying because of their poor health. Second, our study did not consider several potential factors, including socio-economic status and dietary and exercise habits. Lifestyle habits may be a mediator variable, so further studies are needed to confirm the impact of such factors, even though it is difficult to accurately measure them.

## Conclusion

In summary, we found that certain types of occupations substantially influence CKD progression. Further research in a clinical setting is needed to clarify the mechanisms and causal relationships involved.

## Data Availability

Due to the law: Act on the Protection of Personal Information, the data cannot be released to the public. The data will be shared on reasonable request to the corresponding author.

## Acknowledgements

We thank all the staff members in Japan Health Insurance Association.

